# Patients’ perspectives on ecologically sustainable healthcare in general practice

**DOI:** 10.1101/2024.10.18.24310602

**Authors:** E.H. Visser, E.A. Brakema, I.A. Slootweg, H.M.M. Vos, M.A. Adriaanse

**Affiliations:** Department of Public Health and Primary Care, Leiden University Medical Center, Leiden, Netherlands; Department of Health, Medical and Neuropsychology of the Faculty of Social and Behavioural Sciences of Leiden University, Leiden, Netherlands

**Keywords:** sustainable healthcare, climate change, planetary health, patients’ perspective, patient participation, general practice

## Abstract

**Background:** Healthcare contributes substantially to climate change. GPs want to implement sustainable healthcare, but are hesitant; worried that this may jeopardise their patient-physician relationship. However, whether this concern is valid is yet to be assessed.

**Aim:** To explore patients’ perspectives on sustainable healthcare in general practice.

**Design and setting:** In 2022 and 2023 we performed an online study, among Dutch GP patients, using experimental vignettes and a questionnaire.

**Method:** The vignettes described GP appointments for three health complaints with randomly allocated treatment advice, varying in sustainability and explanation, but with comparable health outcomes. The questionnaire assessed patients’ perspectives on sustainable healthcare in general practice. We analysed the vignettes using mixed-design ANOVA and the questionnaire using descriptive statistics and correlations.

**Results:** 801 participants completed the vignettes, and 397 the questionnaire. We found no difference on satisfaction with a doctor’s visit (*P*’s>.238) when comparing a sustainable and a non-sustainable treatment option. The effect of explicitly mentioning sustainability differed per health complaint (dyspnoea: no difference; knee pain: MD=.319, *P*=.002; erythema: MD=-.227, *P*=.003). In the questionnaire, participants reported positive expectations, and trust in the GP and treatment when delivering sustainable healthcare, but were more neutral about the GPs’ role.

**Conclusion:** We found no indication that sustainable treatment advice leads to lower satisfaction with healthcare. The effect of explicitly mentioning sustainability was minimal and differed per health complaint. When directly asked, patients were mainly positive about sustainable healthcare. These results could encourage GPs to introduce sustainable treatment advice, without worrying about negatively influencing patient satisfaction.

**How this fits in:** GPs and other healthcare professionals increasingly want to implement sustainable healthcare, but are hesitant to do so, fearing that it will jeopardise their patient-physician relationship. However, no studies have been conducted to assess how patients actually respond to sustainable healthcare in general practice. In this study among GP patients, we found: no indication that sustainable treatment advice, in scenarios with comparable health outcomes, leads to lower satisfaction with a doctor’s visit; that the effect of explicitly mentioning sustainability on satisfaction with a doctor’s visit had a minimal effect that differed per health complaint; and that patients were mainly positive about sustainable healthcare when reflecting on this topic in a questionnaire. These findings may encourage GPs to introduce sustainable treatment options in their consultations, without worrying about negatively influencing patient satisfaction.

## Introduction

Human activities place the planet’s ecosystems under severe pressure, resulting in climate change and other ecological crises.(1) These crises affect our living environment and thereby increase human morbidity and mortality;(2) the World Health Organization even considers climate change ‘the single biggest health threat facing humanity’.(3) Paradoxically, healthcare contributes substantially to these ecological crises through, for example, resource depletion, (chemical) pollution, and CO2 emissions.(4-8) Therefore, healthcare professionals, including general practitioners (GPs) worldwide,(9-11) urgently call for a transition to ecologically sustainable healthcare.

Fortunately, there is a growing body of knowledge on how to mitigate the environmental footprint of healthcare. This includes, for example, knowledge on how to reduce the environmental harm of surgery,(12) the carbon footprint of different modes of delivery,(13) or the environmental benefit of using dry powder inhalers compared to metered dose inhalers.(14) The latter is even already included in GP guidelines.(15)

However, guidelines are frequently insufficient to change healthcare professional behaviour:(16) knowledge does not automatically translate into motivation to change,(17, 18) let alone, actual behaviour change, as these are dependent on other determinants as well.(19) For providing sustainable healthcare, a key barrier that has been identified is healthcare professionals’ negative expectations about patients’ responses to sustainable healthcare,(20, 21) and corresponding concerns about jeopardizing their patient-physician relationship. However, patients’ responses towards sustainable healthcare have only limitedly been explored and not yet in the context of general practice,(22-24) so whether these concerns are warranted is unclear. Therefore, we aimed to study patients’ responses to sustainable healthcare in general practice and the potential influence on satisfaction with care.

## Methods

We conducted an online study including experimental vignettes and a questionnaire. The study was approved by the Departmental Ethical Review Board of Leiden University Medical Center (#22-3046) and preregistered on AsPredicted (https://aspredicted.org/2df6w.pdf). Data and syntaxes will be made available on the Open Science Framework (https://osf.io/pvd4u/) after publication.

### Study design

The study consisted of two parts, i.e. experimental vignettes with random allocation; and a questionnaire, similar for all participants. We collected data in two waves, wave one in October 2022 and wave two in March 2023. We report data for the experimental vignette study of both waves and the questionnaire data of wave two; as the questionnaire data of wave one were unusable due to a coding error.

See supplementary figure 1 for a study flow diagram.

### Participants

We included a representative sample of adult, Dutch, GP patients, via an online panel (Flycatcher.eu). We obtained digital informed consent prior to participation.

### Materials

#### Baseline demographics

Baseline demographics included age, sex/gender, education level, type of living area, having (grand)children, trust in the GP, number of GP visits and self-rated health status.

#### Experimental vignettes

We did not inform participants that the topic of our study was sustainable healthcare prior to the experimental vignettes, as we aimed to obtain an unbiased response by participants, blind to the other conditions and purpose of the study. The experimental vignettes presented each participant with three scenarios about a GP visit. Each scenario described a different health complaint (i.e. ‘Health complaint’): dyspnoea, knee pain, and erythema. These health complaints have at least two treatment options that have comparable health outcomes, but, with current knowledge, differ in their environmental impact.(15, 25, 26) Participants were randomly assigned to one of four predefined possible doctor’s advices that were then applied to all three scenarios. These advices (i.e. ‘Type of advice’) varied in whether it was sustainable or not, and whether sustainability was named as an argument:

- the *Non-Sustainable Advice*: only the non-sustainable treatment option was offered;
- the *Sustainable Advice*: only the sustainable treatment option was offered;
- the *Sustainable with Alternative Advice*: the sustainable treatment option was offered, while making explicit that there is another option;
- the *Sustainable made Explicit Advice*: the sustainable treatment option was offered, while making explicit that there is another option, with sustainability as an explicit argument for suggesting this advice

See box 1 for an example.

This resulted in a 3x4 design: the three different health complaints assessed differences within subjects; the four types of advice assessed differences between subjects.

You visit your GP because you sometimes feel short of breath. After taking your medical history and performing a physical examination, the GP diagnoses you with a lung disease, such as asthma or COPD. The doctor wants to start you on medication.

There are different types of medication available:

1. a metered dose inhaler/puffer or
2. a dry powder inhaler

He/she recommends a dry powder inhaler to help you feel less short of breath. The GP tells you that this type of medication is better for the environment than the metered dose inhaler.

confidence in the treatment, and the feeling that their health is a priority to the GP (five-point Likert scales; 1: strongly disagree; to 5: strongly agree).

After participants completed the vignettes, we asked them to indicate whether they had experienced similar health complaints and treatment advice.

#### Sustainable Healthcare Questionnaire in General Practice (SHQ_GP)

We developed the SHQ_GP to assess patients’ general perspective on sustainable healthcare within the context of general practice. The questionnaire included 15 items, assessing seven constructs based on broader literature on patient satisfaction and healthcare professional behaviour (five-point Likert scale).

#### Overall opinion on climate change

We administered five items of the ‘single items scale’ of Valkengoed et al. (2021) to assess the overall opinion on climate change (five-point Likert scale).(27)

The experimental vignettes, SHQ-GP and development rationale are available in the Supplementary Data.

### Statistical analysis

We used IBM SPSS Statistics version 29 for statistical analyses. We present the demographic variables using frequencies, percentages, medians, and ranges.

#### Vignettes

We conducted Chi-Square and Kruskall-Wallis tests to check for successful randomization. We assessed the effect of ‘Type of advice’ (between subjects) and ‘Health complaint’ (within subjects) on ‘Satisfaction with the doctor’s visit’ using mixed-design ANOVA, presenting the Huynh-Feldt estimates. We used Spearman’s Rho to explore the relationship between demographics, experience with the diseases, overall opinion on climate change, and ‘Satisfaction with the doctor’s visit’ for each ‘Health complaint’.

#### Questionnaire

We analysed responses to the SHQ_GP using percentages, medians, interquartile ranges (IQR); and Cronbach’s Alpha per SHQ_GP construct. We used Spearman’s Rho to explore the relationship between demographics, overall opinions on climate change, and the SHQ_GP constructs.

For the sake of transparency, adjustments to our preregistration and rationale for methodological choices are available in the Supplementary Data.

## Results

We sent the survey to 1,330 participants, of which 801 (wave one and two) completed the experimental vignettes and 397 (wave two) completed the SHQ_GP.

### Demographics

Participants’ average age was 53 years and sex/gender was equally distributed (50.7%, n=206 males). Participants indicated a mean of two GP visits per year. Demographics are available in table 1.

**Table 1.**
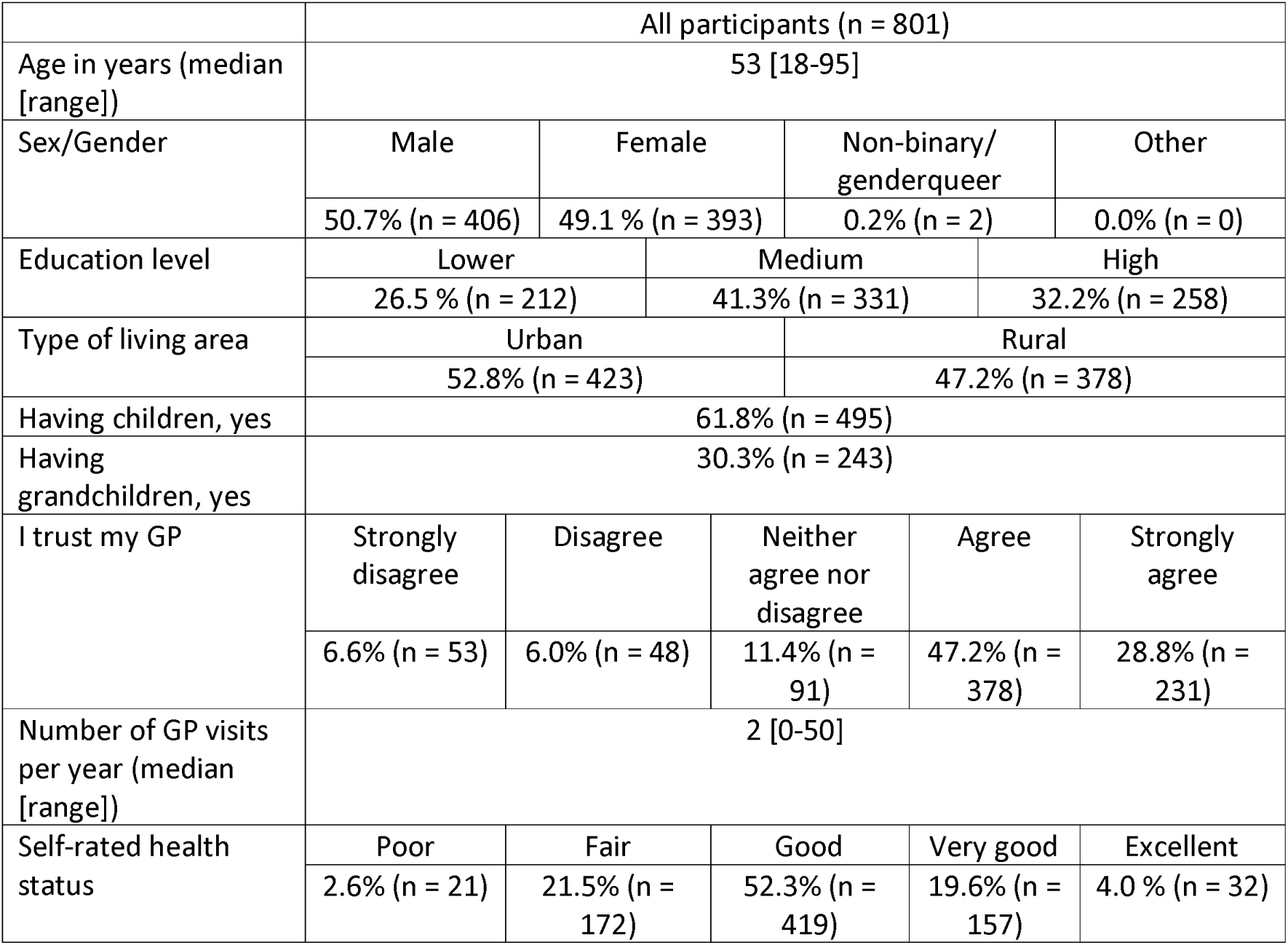
Demographic variables.

### Experimental Vignettes

#### Randomization check

Participants were equally distributed across the types of advice (n=198 to n=204). We found no statistically significant differences between the four groups (type of advice) on demographics, control variables and overall opinion on climate change, except on ‘Number of GP visits per year’ (P=.04). Conform preregistration, we included this variable as covariate in the main analysis, but report the analyses without this covariate as it was not statistically significant.

#### Main analysis

The mixed-design ANOVA on ‘Satisfaction with the doctor’s visit’ did not yield a statistically significant main effect of ‘Type of advice’ (*P*=.41). The within subjects’ effect of ‘Health complaint’ was statistically significant (F (1.91, 1521.52)=326.98, *P*<.001, partial η2=.29), with the highest scores for Erythema (M=3.87, SD=0.77), then for Dyspnoea (M=3.78, SD=0.87) and the lowest score for Knee pain (M=3.12, SD=0.99). The ‘Type of advice’ x ‘Health complaint’ interaction was also statistically significant (F (5.73, 1521.52)=8.89, *P*<.001, partial η2=.03). Pairwise comparisons were conducted to decompose the interaction.

These exploratory pairwise comparisons within each of the ‘Health complaints’ revealed that within Dyspnoea, the Sustainable with Alternative Advice (M=3.90, SD=.85) received statistically significant higher scores on satisfaction than the Non-Sustainable Advice (M=3.70, SD=.88), as well as Sustainable made Explicit Advice (M=3.70, SD=.99). All other *P*s were >.24. Within Knee pain, both the Sustainable with Alternative Advice (M=3.25, SD=.98) and the Sustainable made Explicit Advice (M=3.26, SD=.98) scored statistically significant higher on satisfaction as compared to both the Non-Sustainable Advice (M=3.04, SD=1.01) as well as the Sustainable Advice (M=2.95, SD=.96). All other Ps were >.37. Lastly, within Erythema, the Sustainable made Explicit Advice (M=3.74, SD=.89) scored statistically significant lower than both the Non-Sustainable Advice (M=3.92, SD=.76) and the Sustainable Advice (M=3.97, SD=.64). All other Ps were >.21. All pairwise comparisons are available in supplementary table 1 and figure 1 gives a visual representation with mean scores on ‘Satisfaction with a doctor’s visit’ across the three ‘Health complaints’ and four ‘Types of advice’.

**Figure 1.**
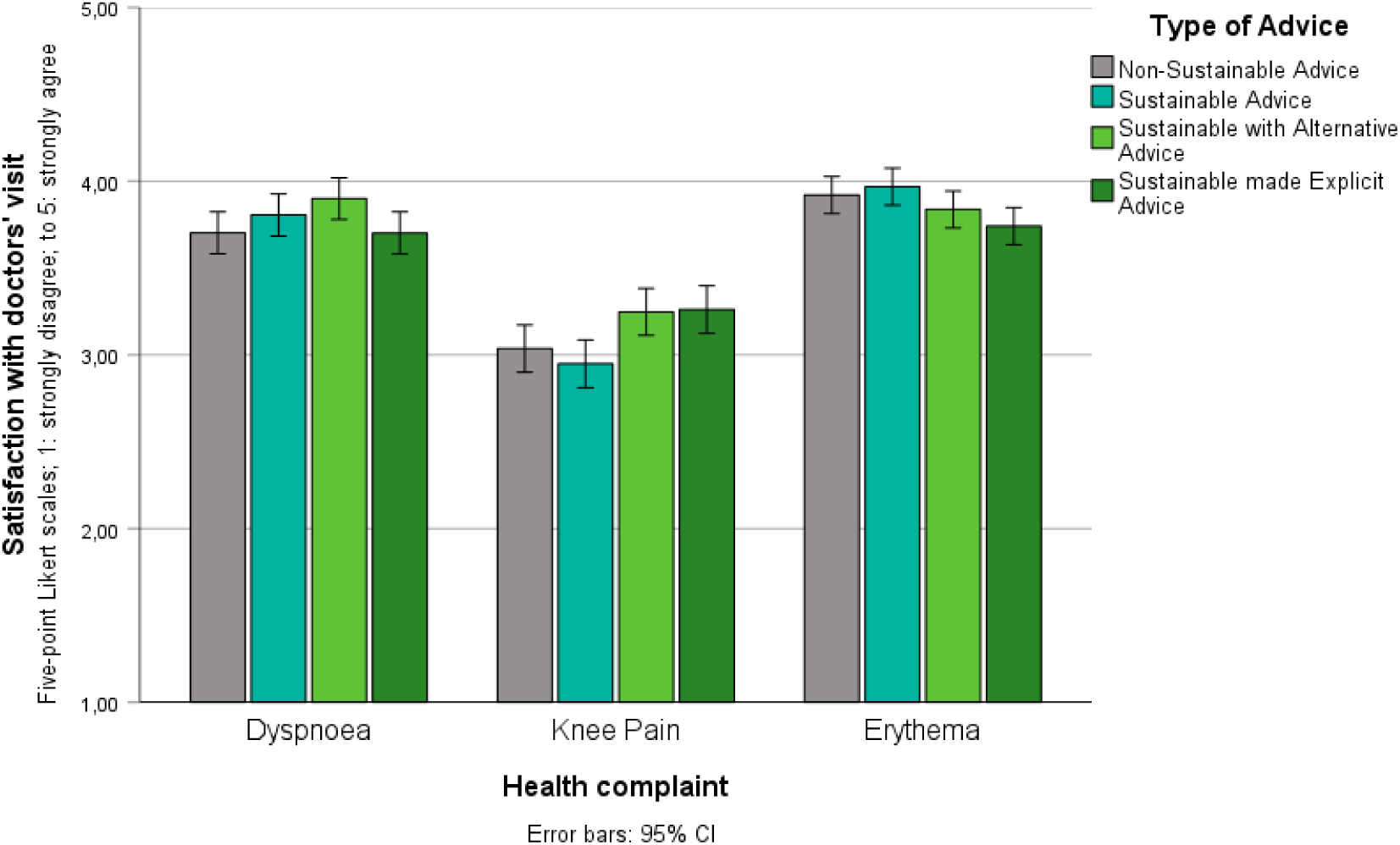
‘Satisfaction with the doctor’s visit’ (five-point Likert scales; 1: strongly disagree; to 5: strongly agree) per ‘Health complaint’, with separate bars per ‘Type of Advice’.

See Supplementary Table 2 for correlation between demographics, overall opinion on climate change and health complaints.

##### Sustainable Healthcare Questionnaire in General Practice (SHQ_GP)

The results of the SHQ-GP show that patients are generally neutral to positive about sustainable healthcare, with low percentages expressing negative opinions to the statements (strongly disagree, or strongly agree in negatively stated items < 6.0%). Overall, patients reported positive expectations, trust in the GP and treatment when delivering sustainable healthcare, but are more neutral about the role or task of the GP in sustainable healthcare. A complete overview can be found in figure 2, showing the percentages per given answer, accompanied by median and IQR per item and Cronbach’s Alpha clustered per construct.

**Figure 2.**
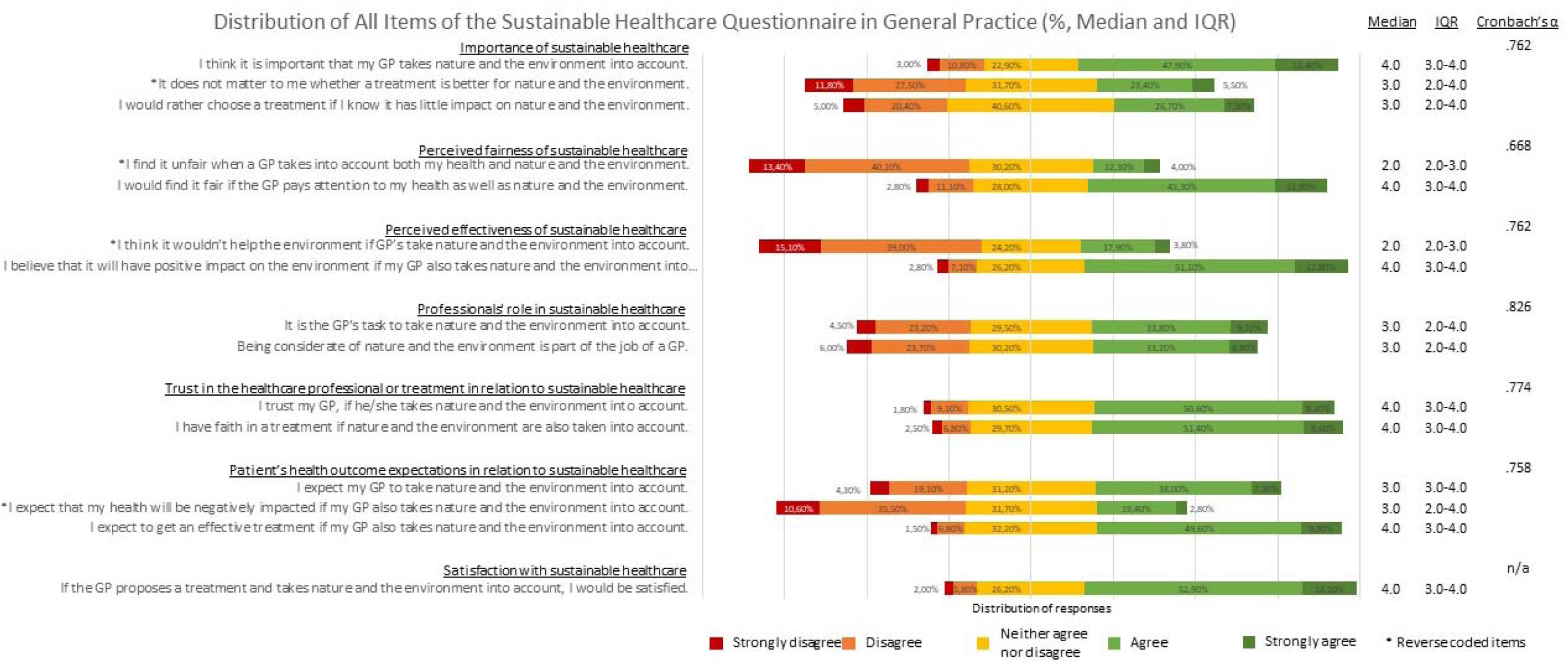
Distribution of scores of all SHQ_GP items clustered per construct. *\* Negatively stated items*

Correlations between demographics and SHQ_GP constructs in table 2 show that age correlated positively with the perceived *role of the GP* in sustainable healthcare. For sex/gender, there is a small correlation indicating that women or non-binary/gender queer people have a more positive stance regarding *importance* and *effectiveness* of sustainable healthcare. Finally, an increase in education level is associated with a more positive stance regarding *fairness* and *effectiveness*. These correlations showed no consistency over the constructs. There is no statistically significant correlation between demographic variables and *trust, expectations,* or *satisfaction with treatment.* See Supplementary Table 3 for the complete correlation table.

**Table 2.** Correlation table of demographic variables (age, sex/gender, education level) with SHQ_GP constructs. *\*\* Correlation is statistically significant at the 0.01 level (2-tailed).* *\* Correlation is statistically significant at the 0.05 level (2-tailed).*

|  |  | Demographics |  |  | Constructs in SHQ_GP |  |  |  |  |  |  |
| --- | --- | --- | --- | --- | --- | --- | --- | --- | --- | --- | --- |
|  |  | Age in years | Sex/Gender | Education level | Importance of sustainable healthcare | Perceived fairness | Perceived effectiveness | Health professional role | Trust | Expectation | Satisfaction treatment |
| Demographics |  |  |  |  |  |  |  |  |  |  |  |
| Age in years | r<sub>s</sub> | -- |  |  |  |  |  |  |  |  |  |
| Sex/gender | r<sub>s</sub> | -,289** | -- |  |  |  |  |  |  |  |  |
| Education level in three categories | r<sub>s</sub> | -,323** | 0,053 | -- |  |  |  |  |  |  |  |
| Constructs in SHQ_GP |  |  |  |  |  |  |  |  |  |  |  |
| Importance of sustainable healthcare | r<sub>s</sub> | 0,066 | ,133** | 0,069 | -- |  |  |  |  |  |  |
| Perceived fairness | r<sub>s</sub> | -0,061 | 0,061 | ,155** | ,684** | -- |  |  |  |  |  |
| Perceived effectiveness | r<sub>s</sub> | -0,09 | ,118* | ,194** | ,714** | ,719** | -- |  |  |  |  |
| Health professional role | r<sub>s</sub> | ,197** | -0,03 | 0,004 | ,714** | ,545** | ,535** | -- |  |  |  |
| Trust | r<sub>s</sub> | -0,003 | -0,037 | 0,076 | ,626** | ,671** | ,616** | ,548** | -- |  |  |
| Expectation | r<sub>s</sub> | 0,095 | 0,009 | 0,051 | ,766** | ,740** | ,732** | ,722** | ,742** | -- |  |
| Satisfaction treatment | r<sub>s</sub> | -0,073 | 0,045 | 0,071 | ,595** | ,646** | ,621** | ,473** | ,687** | ,643** | -- |

## Discussion

### Summary

In this study we explored patients’ perspectives on sustainable healthcare in general practice and the potential influence of giving sustainable treatment advice on satisfaction with care, in scenarios with comparable health outcomes. We found no differences in patient satisfaction when a more sustainable treatment option is advised compared to a less sustainable treatment option. The effect of explicitly naming sustainability as an argument in explaining the choice for the treatment option, as compared to advising the sustainable option without such an explanation on patient satisfaction, was small and differed per health complaint. Lastly, most participants responded relatively positive when asked about sustainable healthcare in general practice in our questionnaire.

### Satisfaction with a doctor’s visit is not negatively – nor positively – influenced by recommending a sustainable treatment option

Regardless of whether participants deliberately reflected on sustainable healthcare in general practice (questionnaire) or when comparing patient responses who were randomly assigned to sustainable versus non-sustainable treatment options with comparable health outcomes in hypothetical GP scenarios, without making the topic of sustainable healthcare explicit (experimental vignettes), sustainable healthcare did not yield negative responses regarding patients’ satisfaction. The concerns about undermining trust, as expressed by Resnik and Pugh,(28) thus do not seem grounded in patient opinion. In fact, our results can be seen as fertile ground to further encourage and support the implementation of sustainable healthcare.

### Explicitly naming sustainable healthcare as an argument to support the decision to prescribe a sustainable treatment option has small, mixed effects across the scenario’s

When explicitly naming sustainability as an argument compared to not naming it explicitly, patient satisfaction differed per health complaint; it was marginally higher for Knee Pain, lower in Erythema, and indifferent in Dyspnoea. Why this difference exists across health complaints is to be determined in future studies. Present findings, leave the ethical question of how much we, as GPs, are willing to trade off in terms of satisfaction and trust when this leads to an increase in awareness regarding the importance of sustainable healthcare, also for the sake of health. Especially since a 2015 study found that primary care physicians are considered the most trusted source when it comes to health information related to global warming, making GPs potentially important stakeholders in raising awareness.(29)

### Patients respond neutral to positively to sustainable healthcare in general

The SHQ_GP showed an overall neutral to positive response from participants towards sustainable healthcare in the context of general practice. This seems to be largely independent of presumed indicators of a higher commitment to ecological crises, such as lower age, female gender or higher education level.(30) Most participants expected to be satisfied with receiving sustainable healthcare, reported positive expectations towards sustainable healthcare, and trust in both the GP and treatment when sustainable healthcare was delivered. Most participants also indicated to perceive sustainable healthcare as important. Notably, there is less consensus regarding the role or task of the GP in the context of sustainable healthcare; participants appear to exhibit a more neutral stance on this matter. Despite the majority having a neutral to positive attitude toward sustainable healthcare, there is also a non-negligible group of people (outspokenly) negative about the importance and expectations regarding sustainable healthcare. This makes it even more important to have situational awareness and to match the treatment advice, and in this case in particular the explanation, to the individual patient.

### Strengths and limitations

We conducted this study combining expertise from general practice, sustainable healthcare, and behavioural science. This allowed us to combine the reality of everyday medical practice, ambitions regarding sustainable healthcare, and aligning these with behavioural science knowledge as well as methodology (e.g., experimental vignettes). In addition, the large and representative sample strengthens our confidence regarding the robustness of our findings. Lastly, the use of experimental vignettes and the absence of advance notification to patients about sustainable healthcare provides the currently closest approximation to the actual situation and reduces the risk of response bias.

As a limitation, our findings rely upon patients’ hypothetical perspectives as no real-life behavioural responses were measured. However, these data are exploratory and pioneering in a new field. Secondly, despite our attempt to use a representative sample, the sample exhibited a lower frequency of doctor visits than the median reported in national databases.(31) This discrepancy may indicate that our sample is healthier than the average population or may be the result of memory bias as this is self-reported data. Still, these findings must be interpreted with caution in patients who visit their GP more frequently. Lastly, we included vignettes with similar patient outcomes and in a non-acute setting. Future research should replicate our findings with different patient outcomes and in other care settings.

### Conclusion

The patients’ perspective on sustainable healthcare in general practice tends to be generally positive. This suggests that we, as GPs, can start changing our behaviour and advise sustainable treatment options with comparable health outcomes, without worrying about negatively influencing patient satisfaction. Explicitly discussing sustainability as an argument to raise awareness should be treated with more care. Future research will have to focus on whether our findings can be extrapolated to other settings or to treatments in which patient outcomes are not comparable.

## Data Availability

Data and syntaxes will be made available on the Open Science Framework after publication

https://osf.io/pvd4u/

## Ethical approval

The study was approved by the Departmental Ethical Review Board of Leiden University Medical Centre (#22-3046).

## Data

The study was preregistered on AsPredicted (https://aspredicted.org/2df6w.pdf). Data and syntaxes will be made available on the Open Science Framework (https://osf.io/pvd4u/) after publication.

## Acknowledgments

The authors would like to thank Petra van Peet (GP), Roeland Watjer (GP trainee) and Marjolein Manders - Schoonakker (GP trainee) for checking if the experimental vignettes were in line with everyday general practice. Furthermore, the authors would like to thank Iris Wichers (senior researcher and GP) and Caroline Moermond (senior researcher) for sharing their knowledge on sustainable healthcare options in the designing phase of the experimental vignettes.

