## Supplementary Data for "Patients’ perspectives on ecologically sustainable healthcare in general practice"

### Supplementary Data - Patients' perspectives on ecologically sustainable healthcare in general practice

### 1. FIGURE S1

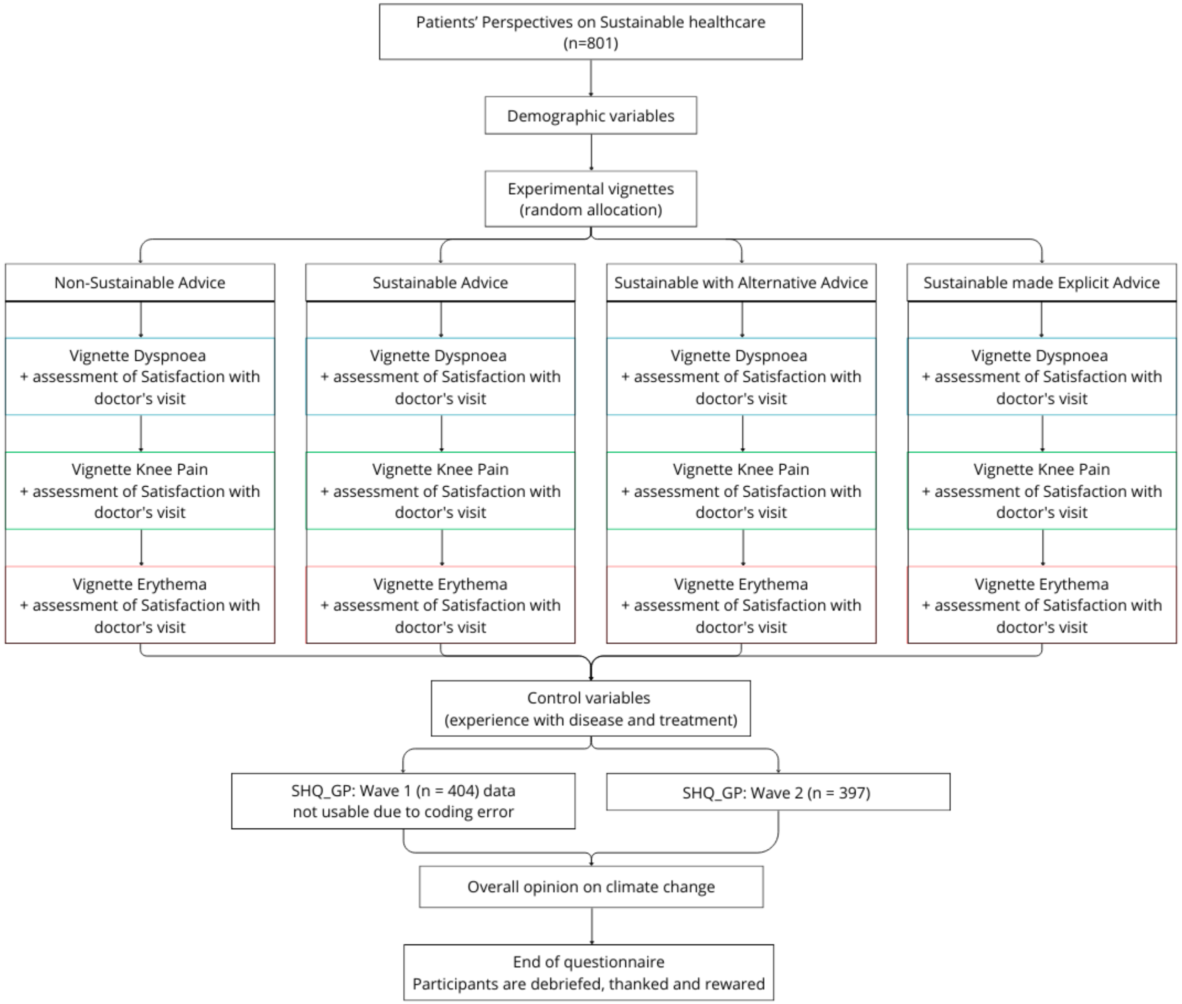

*Figure S1 – Study flow diagram*

#### 2. EXPERIMENTAL VIGNETTES AND SUSTAINABLE HEALTHCARE QUESTIONNAIRE IN GENERAL PRACTICE (SHQ_GP)

#### Translated from Dutch (original) to English

Demographics

- Age:
  - Open
- Sex/gender
  - Male
  - Female
  - Non-binary/gender queer
  - I describe my gender as:
- What is the highest education you have completed? (except for primary education, this means that you have obtained a diploma for this education)
  - No or primary education
  - Pre-vocational secondary education
  - Pre-vocational secondary education / Senior general secondary education until 3rd year/ Pre-university education until 3rd year
  - Secondary vocational education
  - Senior general secondary education/ Pre-university education
  - Higher professional education first year
  - Higher professional education bachelor
  - Higher professional education master
  - University education first year
  - University education bachelor
  - University education master, PhD or post-doctoral
- I live in:
  - A city
  - A village/countryside
- Do you have children?
  - Yes
  - No
- Do you have grandchildren?
  - Yes
  - No

GP visits and health status

- In the past 12 months, how often did you contact your GP?
  - *By this we mean a physical or video call consultation with the GP himself or herself. (If you have had no contact fill in 0)*
  - Open answer field (.... times)
- I trust my GP. (*Some practices have more than one GP. If this is the case with you: please fill in the answer for the GP you have seen most often in the past 12 months*)
  - Strongly disagree
  - Disagree
  - Neither agree nor disagree

- Agree
- Strongly agree

- In general, would you say your health is:
  - Poor
  - Fair
  - Good
  - Very good
  - Excellent

Section A: Vignettes

We would now like to ask you to empathise with three situations, described below. These are situations that describe an appointment with your GP. Try to imagine yourself in the situation described as best you can. Picture yourself as the patient in that situation. Afterwards, we will ask you some questions about what you would think and how you would feel in that situation.

Vignette 1: Dyspnoea

*Condition 1 (Non-Sustainable Advice):*

You visit your GP because you sometimes feel short of breath. After taking your medical history and performing a physical examination, the GP diagnoses you with a lung disease, such as asthma or COPD. The doctor wants to start you on medication. He/she recommends a metered dose inhaler/puffer to help you feel less short of breath.

*Condition 2 (Sustainable Advice):*

You visit your GP because you sometimes feel short of breath. After taking your medical history and performing a physical examination, the GP diagnoses you with a lung disease, such as asthma or COPD. The doctor wants to start you on medication. He/she recommends a dry powder inhaler to help you feel less short of breath.

*Condition 3 (Sustainable with Alternative Advice):*

You visit your GP because you sometimes feel short of breath. After taking your medical history and performing a physical examination, the GP diagnoses you with a lung disease, such as asthma or COPD. The doctor wants to start you on medication.

There are different types of medication available:

1. a metered dose inhaler/puffer or
2. a dry powder inhaler

He/she recommends a dry powder inhaler to help you feel less short of breath.

*Condition 4 (Sustainable made Explicit Advice):*

You visit your GP because you sometimes feel short of breath. After taking your medical history and performing a physical examination, the GP diagnoses you with a lung disease, such as asthma or COPD. The doctor wants to start you on medication.

There are different types of medication available:

3. a metered dose inhaler/puffer or
4. a dry powder inhaler

He/she recommends a dry powder inhaler to help you feel less short of breath. The GP tells you that this type of medication is better for the environment than the metered dose inhaler.

Vignette 2: Knee Pain

*Condition 1 (Non-Sustainable Advice):*

You visit your GP because your right knee is painful when walking. After taking your medical history and performing a physical examination, the GP diagnoses osteoarthritis of the knee. The doctor wants to start treatment for this. He/she recommends that you take paracetamol and optionally diclofenac (stronger painkiller) to reduce the pain.

*Condition 2 (Sustainable Advice):*

You visit your GP because your right knee is painful when walking. After taking your medical history and performing a physical examination, the GP diagnoses osteoarthritis of the knee. The doctor wants to start treatment for this. He/she recommends that you take paracetamol to reduce the pain.

*Condition 3 (Sustainable with Alternative Advice):*

You visit your GP because your right knee is painful when walking. After taking your medical history and performing a physical examination, the GP diagnoses osteoarthritis of the knee. The doctor wants to start treatment for this. There are several options:

1. Paracetamol or
2. Paracetamol and optionally diclofenac (stronger painkiller).

He/she recommends that you take paracetamol to reduce the pain.

*Condition 4 (Sustainable made Explicit Advice):*

You visit your GP because your right knee is painful when walking. After taking your medical history and performing a physical examination, the GP diagnoses osteoarthritis of the knee. The doctor wants to start treatment for this. There are several options:

1. Paracetamol or
2. Paracetamol and optionally diclofenac (stronger painkiller).

He/she recommends that you take paracetamol to reduce the pain. The GP tells you that paracetamol is less environmentally damaging than diclofenac (stronger painkiller).

Vignette 3: Erythema

*Condition 1 (Non-Sustainable Advice):*

You visit your GP because you have a redness on your left lower leg. After taking your medical history and performing a physical examination, the GP diagnoses you with a skin infection. The doctor wants to give you medication for this. He/she recommends the antibiotic clarithromycin so that your skin can recover. You take this medicine twice a day and the course of treatment lasts for 10 days.

*Condition 2 (Sustainable Advice):*

You visit your GP because you have a redness on your left lower leg. After taking your medical history and performing a physical examination, the GP diagnoses you with a skin infection. The doctor wants to give you medication for this. He/she recommends the antibiotic flucloxacillin so that your skin can recover. You take this medicine 4 times a day and the course of treatment lasts for 10 days.

*Condition 3 (Sustainable with Alternative Advice):*

You visit your GP because you have a redness on your left lower leg. After taking your medical history and performing a physical examination, the GP diagnoses you with a skin infection. The doctor wants to give you medication for this. Several types of antibiotics are available:

1. Clarithromycin, you take this medicine twice a day and the course of treatment lasts for 10 days.
2. Flucloxacillin, you take this medicine 4 times a day and the course of treatment lasts 10 days.

He/she recommends the antibiotic flucloxacillin so that your skin can recover. You take this medicine 4 times a day and the course of treatment lasts for 10 days.

*Condition 4 (Sustainable made Explicit Advice):*

You visit your GP because you have a redness on your left lower leg. After taking your medical history and performing a physical examination, the GP diagnoses you with a skin infection. The doctor wants to give you medication for this. Several types of antibiotics are available:

1. Clarithromycin, you take this medicine twice a day and the course of treatment lasts for 10 days.
2. Flucloxacillin, you take this medicine 4 times a day and the course of treatment lasts 10 days.

He/she recommends the antibiotic flucloxacillin so that your skin can recover. You take this medicine 4 times a day and the course of treatment lasts for 10 days. The GP tells you that flucloxacillin is less harmful to the environment than clarithromycin.

*Statements accompanying each vignette*

*(5-point Likert scale: Completely disagree; Disagree; Neither agree nor disagree; Agree; Completely agree)*

The statements below are about the scenario you have just read. Please keep in mind that you are the patient in the story. You can indicate for each statement whether you agree or disagree.

- I am satisfied with this treatment.
- I trust this GP.
- I have confidence in this treatment.
- This GP pays attention to my health.

- What question would you like to ask the GP after this explanation?
    - Open field.

Diseases and treatments in the vignettes

- Have you ever had lung disease?
    - If yes:
        - Do you have lung disease now?
        - Have you had lung disease in the past?
        - Have you ever taken medication for shortness of breath from lung disease?
            - If yes,
                - Was this a metered dose inhaler/puffer?
                    - Yes
                    - No
                    - I don't know
                - Was this a dry powder inhaler?
                    - Yes
                    - No
                    - I don't know
            - If yes: Were you satisfied with the treatment?
- Have you ever had knee problems?
    - If yes
        - Do you have knee problems now?
        - Did you have knee problems in the past?
        - Have you ever taken medication for the pain of knee pain?
            - If yes
                - Was this paracetamol?
                    - Yes
                    - No
                    - I don't know
                - Was this diclofenac?
                    - Yes
                    - No
                    - I don't know
        - If yes: Were you satisfied with the treatment?
- Have you ever had a skin infection?

- If yes
  - Do you currently have a skin infection?
  - Did you have a skin infection in the past?
  - Have you ever taken medication for a skin infection?
    - If yes
      - Was this flucloxacillin?
        - Yes
        - No
        - I don't know
      - Was this clarithromycin?
        - Yes
        - No
        - I don't know
    - If yes: Were you satisfied with the treatment?

<u>Section B</u>: Opinion on sustainable healthcare

*(5-point Likert scale: completely disagree, disagree, disagree not disagree, agree, agree, completely agree)*

Healthcare (*e.g. GPs, hospitals, and other healthcare providers*) has an impact on nature and the environment. For example, with the energy consumed, the waste produced, and the chemicals from medication that end up in the water via urine. To reduce that impact, more and more healthcare providers are trying to provide 'sustainable healthcare'. This is healthcare that has as little impact on nature and the environment as possible. Below are several statements about healthcare and its impact on nature and the environment. For each statement, you can indicate whether you agree or disagree.

1. I think it is important that my GP takes nature and the environment into account.
2. It is the GP's task to take nature and the environment into account.
3. I find it unfair when a GP takes into account both my health and nature and the environment.
4. I trust my GP, if he/she takes nature and the environment into account.
5. I expect my GP to take nature and the environment into account.
6. It does not matter to me whether a treatment is better for nature and the environment.
7. I think it wouldn't help the environment if GP's take nature and the environment into account.
8. I expect that my health will suffer if my GP also takes nature and the environment into account.
9. Being considerate of nature and the environment is part of the job of a GP.
10. If the GP proposes a treatment and takes nature and the environment into account, I would be satisfied.
11. I have confidence in a treatment if nature and the environment are also taken into account.
12. I would find it fair if the GP pays attention to my health as well as nature and the environment.
13. I would rather choose a treatment if I knew it has little impact on nature and the environment.

14. I believe that it will have positive consequences for the environment if my GP also takes nature and the environment into account.
15. I expect to get an effective treatment if my GP also takes nature and the environment into account.

Opinion about climate change

*(5-point Likert scale: completely agree, agree, disagree not disagree, disagree, completely disagree)*

Below you will see one or more statements about climate change. For each statement you can indicate whether you agree or disagree.

1. I believe that climate change is real.
    a. If no is answered here, the statements below will also be left blank
2. Human activities are a major cause of climate change.
3. Climate change will bring about serious negative consequences
4. My local area will be influenced by climate change.
5. It will be a long time before the consequences of climate change are felt.

<u>End of questionnaire</u>

*Debriefing*:

You have reached the end of the study. We would like to thank you for your participation!

In this study, we are interested in your opinion about sustainable/green healthcare and whether information about sustainable/green healthcare affects how you experience treatment. Sustainable or green healthcare is healthcare provided where there is as little negative impact on nature and the environment as possible. To investigate this, all participants were divided into four groups.

1. The first group was given three scenarios with a non-sustainable option and treatment advice

2. The second group was given three scenarios with a sustainable option and treatment advice

3. The third group was given three scenarios with both the sustainable and non-sustainable option, in which the sustainable option was advised

4. The fourth group was given three scenarios in which both the sustainable and non-sustainable option were presented, in which the sustainable option was advised with explanation that it is better for the environment.

Besides the situations, you also completed general questions about yourself, your GP visits and health, sustainable/green healthcare, and climate change. We drafted these questions because these factors may influence your opinion.

We are announcing the theme of the questionnaire (sustainable care) to you now. We did not announce it at the beginning of the questionnaire because then there is the possibility that some of the participants will give different answers than what they really think.

With this survey, we hope to answer the question of what patients think about sustainable/green healthcare and whether information about sustainable/green healthcare affects how treatment is perceived.

If you have any further questions about this study, please email xxx.

If you wish to withdraw the consent you gave in the informed consent and do not want your data to be used for the above purposes, please tick this box:

□ I want to withdraw the consent I gave and do not want my data to be used.

Thank you again for your participation!

#### Development rationale

This survey is newly constructed for this study because no survey on this topic was available.

The experimental vignettes and SHQ_GP are in level B1 Dutch[a] as much as possible, to make the study accessible for the largest part of the Dutch population.

The total study consists of a minimum of 43 and maximum of 65 questions or statements and three experimental vignettes. The number of questions/statements depends on the answers of the participant.

The survey is divided into two parts:

A. Experimental vignettes
B. Sustainable healthcare questionnaire in general practice (SHQ_GP)

This order within the survey was chosen to minimize the bias of previous questions. The different parts of the questionnaire will be explained in more detail below.

Structure of the survey

The survey starts with baseline demographics (nine questions).

*Baseline demographics*

The following participant variables could potentially influence outcomes on the questionnaire as well as the outcomes of the experimental vignettes.

1. Age
2. Sex/Gender
3. Educational level
4. Type of living area (urban or rural)

Statistics Netherlands (*Dutch: Centraal Bureau voor de Statistiek)* found that: 'It is mainly highly educated people, young people, women and city dwellers who see climate change as a major problem, are concerned about it and consider climate policy by the government to be important.' For this reason age, sex/gender, educational level and type of living area are included as variables in the questionnaire.(1)

5. Having children
6. Having grand children

People who are engaged in environmental sustainability often indicate that they do so in order to be able to properly pass on the planet to the next generation. For this reason, having children or grandchildren might influence the way they perceive sustainable healthcare.

7. Number of GP visits

---

[a] Language level B1 refers to simple Dutch. The vast majority of the population understands texts at language level B1. A text at B1 level consists of easy words that almost everyone uses. And from short, simple, and active sentences. ( https://www.communicatierijk.nl/vakkennis/rijkswebsites/aanbevolen-richtlijnen/taalniveau-b1 )

8. The patient's self-rated health.

For previous experiences with a GP and self-rated health status may influence the way a participant receives a treatment within the vignettes, this is taken into account by asking the number of GP visits and the self-rated health status.

Self-rated health is operationalized as the score on the general perceptions scale of the Short-Form 36 (2). In 1998, a Dutch version of the SF-36 was validated. (3)

9. Trust in the GP

Trust in the GP is one of the outcomes within the experimental vignettes. To enable to possibly correct for an extreme value in trust in the GP prior to the vignettes, a question of trust in the GP is added and measured on a 5-point Likert scale.

*Experimental vignettes*

The participants are presented with three vignettes, with five statements/questions each.

<u>Health problems within the vignettes</u>

The first two experimental vignettes are chosen based on the two guidelines of the Nederlands Huisartsen Genootschap (NHG; Dutch College of General Practitioners), which already contain a link to sustainable healthcare. These guidelines are:

- Guideline Asthma in adults (*Dutch: NHG-richtlijn Astma bij volwassenen versie juli 2020*)
- Guideline Pain (*Dutch: NHG-richtlijn Pijn versie november 2021)*; combined with the Guideline Non-traumatic knee problems (*Dutch: NHG-richtlijn Niet traumatische knieklachten, oktober 2020)*

As these guidelines already contain a link to sustainable healthcare, we refer to these guidelines for more detailed information on this link.

The third experimental vignette is set up in consultation with the NHG and based on the NHG guideline Bacterial skin infections (*Dutch: NHG-richtlijn Bacteriële huidinfecties versie mei 2019*) and information from the Dutch National Institute for Public Health and the Environment (RIVM; Rijksinstituut voor Volksgezondheid en Milieu). The NHG guideline advises the following treatment for a bacterial skin infection:

- First choice of therapy: Flucloxacillin 4 dd 500 mg for 10-14 days
- In case of hypersensitivity to penicillin: clarithromycin 2 dd 500 mg for 10-14 days

Because clarithromycin only has to be taken twice a day and flucloxacillin four times a day, some GPs opt for clarithromycin. However, research by the RIVM shows that the risk quotient for clarithromycin sometimes is higher than one.(4) This means that there is a risk to the water environment. For flucloxacillin this number is not clear yet in the Netherlands. Therefore, clarithromycin is not only the second-choice type of treatment, but also has a potentially higher environmental impact.

We chose these three vignette types for the treatments generally have comparable health outcomes; however, they differ in their impact on the environment or climate. It is important to note that these are examples which, because there is still little scientific evidence on the environmental impact of different treatments, serve as the best possible example but may be invalidated as a sustainable option by future research. This does not affect the outcome measures of the experimental vignettes (the influence of information about sustainable healthcare on the way the treatment is received).

Before reading a vignette, the participants will be asked to imagine themselves to be the patient in the vignette.

<u>Dependent variables</u>

There are four dependent variables: acceptability of the treatment, trust in the GP, confidence in the treatment and the feeling that your health as a patient is a priority for the GP. These four dependent variables were chosen because of their hypothesized importance for current compliance and future doctor visits and treatments.

- Acceptability:
  Sekhon et al. defined acceptability as: 'a multi-faceted construct that reflects the extent to which people delivering or receiving a healthcare intervention consider it to be appropriate, based on anticipated or experienced cognitive and emotional responses to the intervention.' They developed the theoretical framework of acceptability (TFA), which consists of seven component constructs: affective attitude, burden, perceived effectiveness, ethicality, intervention coherence, opportunity costs, and self-efficacy.(5) For this study we chose to measure acceptability using the first construct: affective attitude (how an individual feels about the intervention). We chose this construct to stay as close as possible to how the patient receives a treatment and at the same time keep the survey concise. This will be measured using one item on a 5-point Likert scale.
    - "I am satisfied with this treatment"
- Trust in the GP:
  This dependent variable will be measured using one item on a 5-point Likert scale.
    - "I trust this GP."
- Confidence in the treatment:
  This dependent variable will be measured using one item on a 5-point Likert scale.
- "I have confidence in this treatment." The feeling that your health as a patient is a priority for the GP:
  This dependent variable will be measured using one item on a 5-point Likert scale.
    - "This GP pays attention to my health."

After the four statements a question is added to explore the most important question a patient might have for their GP after the treatment is proposed. This question is added to explore whether informing a patient about the sustainable healthcare considerations of the GP influences the questions they might have for the GP.

- What question would you like to ask the GP after this explanation?

<u>Conditions</u>

Of all vignettes four different conditions (Types of advice) will be available.

1. The first being the non-environmentally sustainable option (the *non-sustainable advice*);
2. The second being the environmentally sustainable option (the *Sustainable advice*);
3. The third being the environmentally sustainable option while informing the patient what the other option is (the *Sustainable with Alternative Advice*);
4. The fourth being the environmentally sustainable option while informing the patient what the other option is, including the explanation that this is environmentally more sustainable (the *Sustainable made explicit advice*).

To be able to compare the vignettes as accurately as possible, we chose to have the vignettes differ as little as possible. Therefore, the first sentences per vignette are identical. In each succeeding condition a word or a small piece of text is changed or added. In this way, possible differences between the outcome measures between the different groups will most likely be based on the effect of information about sustainable healthcare on the way care is received by the patient and not on other changes in parts of the vignette.

The vignettes are checked by independent GPs (trainees) to get as close as possible to the real situation in general practice.

Every participant will be randomly allocated to one of the experimental conditions and will see the three different vignettes of that condition.

This results in a 3 x 4 design as shown in the study flow diagram Figure S1.

<u>Post manipulation questionnaire: Questions on the diseases and treatments in the vignettes</u>

After the experimental vignettes questions are asked about the participants' previous experience with the diseases (pulmonary diseases, knee pain and bacterial skin infections) and respective treatments in the vignettes, for this may give bias in the results of the dependent variables of our experimental vignettes. These are close ended questions, with answer options: Yes / No / I don't know

**Part B: Questionnaire study on sustainable healthcare (minimum of 16 statements, maximum of 20)**

*Sustainable healthcare questionnaire in general practice (SHQ_GP) (15 statements)*

The questions/statements on the patient general perspective on sustainable healthcare were newly constructed for this study, as no standardized or validated questionnaire was available on this topic. Based on literature from social sciences, we defined seven different constructs underpinning the patient's general perspective on sustainable healthcare (figure S2):

1. Importance of sustainable healthcare in healthcare choices (item 1, 6, 13)
2. Perceived fairness of sustainable healthcare(6) (item 3, 12)
3. Perceived effectiveness of sustainable healthcare(7, 8) (item 7, 14)
4. Role of the healthcare professional in sustainable healthcare(9) (item 2, 9)
5. Trust in the healthcare professional or treatment in relation to sustainable healthcare(9) (item 4, 11)
6. Patient's expectations in relation to sustainable healthcare(9) (item 5, 8, 15)
7. Satisfaction with a treatment in relation to sustainable healthcare(10) (item 10)

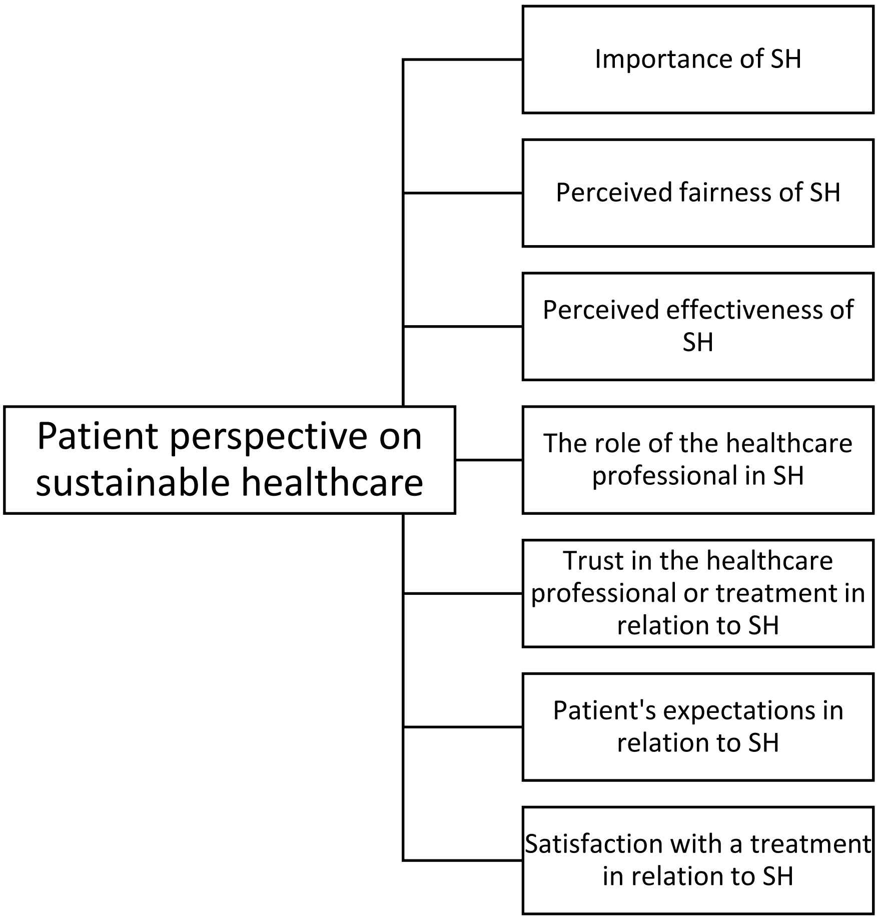

*Figure S2 – Constructs underpinning the statements in the SHQ_GP*

For each construct one, two or three statements/items were established. To keep the questionnaire accessible to as many groups as possible, we chose to use a 5-point Likert scale. Because with an increasing number of options, the accessibility of a questionnaire decreases for participants with a lower educational level. To ensure that similar topics are used throughout the questionnaire and to ensure that the participants can relate to the statements, we chose to ask about the perspective on sustainable healthcare by the GP, rather than by a general healthcare professional. Prior to the statements a brief explanation about sustainable healthcare is given to the participants.

Statements:

1. I think it is important that my GP takes nature and the environment into account.
2. It is the GP's task to take nature and the environment into account.
3. I find it unfair when a GP takes into account both my health and nature and the environment.
4. I trust my GP, if he/she takes nature and the environment into account.
5. I expect my GP to take nature and the environment into account.
6. It does not matter to me whether a treatment is better for nature and the environment.
7. I think it wouldn't help the environment if GP's take nature and the environment into account.
8. I expect that my health will suffer if my GP also takes nature and the environment into account.
9. Being considerate of nature and the environment is part of the job of a GP.

10. If the GP proposes a treatment and takes nature and the environment into account, I would be satisfied.
11. I have confidence in a treatment if nature and the environment are also taken into account.
12. I would find it fair if the GP pays attention to my health as well as nature and the environment.
13. I would rather choose a treatment if I knew it has little impact on nature and the environment.
14. I believe that it will have positive consequences for the environment if my GP also takes nature and the environment into account.
15. I expect to get an effective treatment if my GP also takes nature and the environment into account.

As the statements may appear alike when presented all at once, we chose to show the statements one by one, to avoid confusion.

**Questions on overall opinion on climate change (minimum of one statement, maximum of five statements)**

The participant's overall opinion on climate change is assessed using the single items scale of Van Valkengoed et al. (11) These items are originally on a 7-point Likert scale. To make our questionnaire as accessible as possible for all participants, we chose to convert this to a 5-point Likert scale.

<u>Deception</u>

This survey used deception by not informing participants about the aim of the study before the survey is completed. We chose to not inform participants beforehand, for if a participant knows that the vignettes/questions/statements can be about sustainable healthcare this may influence their answers and the decision whether to participate in the study or not. This influence may be in different ways, e.g.: when the participant is environmentally sustainable active and is more eager to participate or when the participant is in some form against environmental sustainability and is less willing to participate, etc. Because this creates the possibility that our sample is no longer representative for the population, or that possible socially desirable answers will be given, we chose to apply deception.

In the debriefing the set up and aim of the study are explained and there is an op-out option included.

### 3. ADJUSTMENTS TO PREREGISTRATION AND RATIONALE TO METHODOLOGICAL CHOICES

For the sake of transparency, in this section we describe some adjustments to our pre-registration and the rationale behind several of our methodological choices.

- *Data collection*: We would like to note that we initially intended to run one wave of data collection. However, in this initial wave, the SHQ_GP could not be used due to a coding error. For this reason, only the first part considering the experimental vignettes could be used. A second, completely independent wave with a new sample, was conducted after the coding error was solved, that again included the experimental vignettes as well as the SHQ_GP.
- *Statistical analyses*:
    - Experimental vignettes
        - In our preregistration we described that all four dependent variables of the experimental vignettes would be considered as separate dependent variables. However, these four items were found to be highly intercorrelated and to yield similar results when analysed separately. Cronbach’s Alpha was satisfactory for all three ‘Health complaints’; .955 (Dyspnoea), .935 (Knee pain) and .953 (Erythema). Therefore, to increase readability and comprehensibility, we have chosen to combine these four items into one dependent variable: 'satisfaction with the doctor's visit.' And report results for this combined score in this manuscript. The results for the separate items are available in the OSF dataset.
        - For the correlation tables (table S2) we applied no correction because of the exploratory character.
        - After the experimental vignettes we included an open-ended question for exploratory purposes, asking participants to indicate the most important question they had for the GP. To maintain conciseness in this manuscript, we have omitted the results of the open-ended questions from this manuscript. These are available in the OSF dataset.
    - SHQ_GP
        - Conform preregistration, the negatively stated items (3, 6, 7 and 8) were reverse coded and to assess internal consistency of the questionnaire the overall Cronbach’s Alpha and Cronbach’s Alpha per construct were calculated. These are available at the OSF.
        - For the correlation table (table S3) we applied no correction because of the exploratory character.

#### 4. TABLE S1

| Pairwise comparisons | | | | | | | |
|---|---|---|---|---|---|---|---|
| Health Complaint | Type of advice | | Mean Difference (I-J) | Std. Error | Sig. | 95% Confidence Interval for Difference | |
| | | | | | | Lower Bound | Upper Bound |
| Dyspnoea | Non-Sustainable Advice | Sustainable Advice | -,103 | ,087 | ,238 | -,274 | ,068 |
| | | Sustainable with Alternative Advice | -,197* | ,086 | ,023 | -,366 | -,027 |
| | | Sustainable made explicit Advice | ,001 | ,087 | ,993 | -,170 | ,171 |
| | Sustainable Advice | Sustainable with Alternative Advice | -,094 | ,087 | ,279 | -,264 | ,076 |
| | | Sustainable made explicit Advice | ,104 | ,087 | ,236 | -,068 | ,275 |
| | Sustain able with | Sustainable made explicit Advice | ,197* | ,087 | ,023 | ,027 | ,368 |
| Knee Pain | Non-sustainable Advice | Sustainable Advice | ,088 | ,098 | ,372 | -,105 | ,281 |
| | | Sustainable with Alternative Advice | -,211* | ,098 | ,031 | -,403 | -,020 |
| | | Sustainable made explicit Advice | -,225* | ,098 | ,022 | -,418 | -,032 |
| | Sustainable Advice | Sustainable with Alternative Advice | -,299* | ,098 | ,002 | -,492 | -,107 |
| | | Sustainable made explicit Advice | -,313* | ,099 | ,002 | -,507 | -,119 |
| | Sustain able with | Sustainable made explicit Advice | -,014 | ,098 | ,888 | -,206 | ,179 |
| Erythema | Non-Sustainable Advice | Sustainable Advice | -,048 | ,077 | ,532 | -,199 | ,103 |
| | | Sustainable with Alternative Advice | ,083 | ,076 | ,275 | -,066 | ,233 |
| | | Sustainable made explicit Advice | ,179* | ,077 | ,020 | ,028 | ,330 |

| | | | | | | | |
|---|---|---|---|---|---|---|---|
| | Sustainable Advice | Sustainable with Alternative Advice | ,131 | ,077 | ,087 | -,019 | ,282 |
| | | Sustainable made explicit Advice | ,227* | ,077 | ,003 | ,076 | ,379 |
| | Sustain able with | Sustainable made explicit Advice | ,096 | ,077 | ,211 | -,054 | ,246 |

*Table S1 – Pairwise comparisons between different 'Types of advice' per 'Health complaint'.*

** The mean difference is statistically significant at the .05 level.*

### 5. TABLE S2

| | | Demographics | | | | | | | | | Experience with diseases | | | Overall Opinion on Climate Change | | | | | Satisfaction with doctor's visit | | |
|---|---|---|---|---|---|---|---|---|---|---|---|---|---|---|---|---|---|---|---|---|---|
| | | Age in years | Sex/Gender | Type of living area | Education level | Having children | Having grandchildren | Number of GP visits last 12 months | Trust in the GP | Self-rated health status | Experience pulmonary disease | Experience Knee pain | Experience Bacterial skin | I believe that climate change is real. | The main causes of climate change are human activities. | Climate change will bring about serious negative | My local area will be influenced by climate change. | It will be a long time before the consequences of | V1 – Dyspnoea | V2 – Knee Pain | V3 - Erythema |
| **Demographics** | | | | | | | | | | | | | | | | | | | | | |
| Age in years | rs | -- | | | | | | | | | | | | | | | | | | | |
| Sex/Gender | rs | -,257** | -- | | | | | | | | | | | | | | | | | | |
| Type of living area | rs | 0,032 | -0,014 | -- | | | | | | | | | | | | | | | | | |
| Education level | rs | -,298** | ,095** | -0,036 | -- | | | | | | | | | | | | | | | | |
| Having children | rs | -,490** | ,111** | -,126** | ,125** | -- | | | | | | | | | | | | | | | |
| Having grandchildren | rs | -,671** | ,163** | -0,040 | ,247** | ,502** | -- | | | | | | | | | | | | | | |
| Number of GP visits last 12 months | rs | ,115** | ,084* | 0,008 | -0,069 | -0,036 | -,104** | -- | | | | | | | | | | | | | |
| Trust in the GP | rs | 0,037 | -,118** | -0,037 | ,077* | -0,007 | -0,057 | ,079* | -- | | | | | | | | | | | | |
| Self-rated health status | rs | -,244** | -,091* | 0,032 | ,204** | ,075* | ,175** | -,282** | ,109** | -- | | | | | | | | | | | |

| | | | | | | | | | | | | | | | | | |
|---|---|---|---|---|---|---|---|---|---|---|---|---|---|---|---|---|---|
| **Experience with disease** | | | | | | | | | | | | | | | | | |
| Experience pulmonary disease | $r_s$ | -,111** | -,071* | 0,022 | -0,006 | 0,035 | ,096** | -,145** | -0,001 | ,242** | -- | | | | | | |
| Experience Knee pain | $r_s$ | -,092** | 0,007 | 0,027 | 0,051 | ,075* | 0,054 | -,104** | 0,017 | 0,044 | 0,043 | -- | | | | | |
| Experience Bacterial skin infection | $r_s$ | -0,019 | -0,024 | -0,012 | -0,032 | 0,014 | -0,031 | -,138** | -0,033 | ,090* | ,088* | 0,058 | -- | | | | |
| **Overall opinion on Climate Change** | | | | | | | | | | | | | | | | | |
| I believe that climate change is real. | $r_s$ | -,162** | 0,062 | -,087* | ,266** | ,134** | ,089* | -0,027 | ,108** | ,095** | -0,018 | 0,012 | -0,037 | -- | | | |
| The main causes of climate change are human activities. | $r_s$ | -,159** | ,073* | -,075* | ,207** | ,144** | ,109** | 0,003 | ,112** | ,128** | 0,032 | 0,030 | -0,032 | ,580** | -- | | |
| Climate change will bring about serious negative consequences. | $r_s$ | -,131** | ,136** | -0,060 | ,243** | ,155** | ,092* | 0,018 | ,106** | 0,055 | -0,031 | 0,022 | -0,053 | ,644** | ,672** | -- | |
| My local area will be influenced by climate change. | $r_s$ | -,085* | ,125** | -0,023 | ,215** | 0,065 | ,071* | 0,000 | 0,058 | 0,015 | -0,039 | 0,033 | -0,032 | ,557** | ,534** | ,705** | -- |

| | | | | | | | | | | | | | | | | | | | | | |
|---|---|---|---|---|---|---|---|---|---|---|---|---|---|---|---|---|---|---|---|---|---|
| It will be a long time before the consequences of climate change are felt. | $r_s$ | ,087* | -,131** | 0,022 | -,160** | -,086* | -0,060 | ,088* | -0,029 | -0,016 | ,072* | -0,017 | 0,038 | -,507** | -,417** | -,494** | -,491** | -- | | | |
| **Satisfaction with doctor's visit** | | | | | | | | | | | | | | | | | | | | | |
| Vignette 1 - Dyspnoea | $r_s$ | 0,030 | -,110** | 0,058 | -0,057 | -0,021 | -0,060 | 0,037 | ,297** | 0,055 | 0,003 | 0,061 | 0,004 | 0,048 | 0,067 | 0,061 | 0,022 | 0,032 | -- | | |
| Vignette 2 – Knee Pain | $r_s$ | ,193** | -,086* | 0,032 | -,129** | -0,050 | -,148** | ,103** | ,229** | -0,001 | 0,026 | 0,053 | 0,035 | -,078* | -0,032 | -0,027 | -0,057 | ,085* | ,452** | -- | |
| Vignette 3 - Erythema | $r_s$ | 0,061 | -,128** | ,072* | -0,034 | -0,007 | -,082* | 0,030 | ,313** | ,098** | 0,027 | 0,034 | -0,018 | 0,053 | 0,044 | ,080* | 0,040 | -0,017 | ,568** | ,421** | -- |

*Table S2 – Correlation table for demographics, overall opinion on climate change and health complaints using Spearman's' Rho*

*** Correlation is significant at the 0.01 level (2-tailed).*

** Correlation is significant at the 0.05 level (2-tailed).*

### 6. TABLE S3

| | | Demographics | | | | | | | | | Overall Opinion on Climate Change | | | | | Constructs in SHQ_GP | | | | | | |
|---|---|---|---|---|---|---|---|---|---|---|---|---|---|---|---|---|---|---|---|---|---|---|
| | | Age in years | Sex/Gender | Type of living area | Education level | Having children | Having grandchildren | Number of GP visits last 12 months | Trust in the GP | Self-rated health status | I believe that climate change is real. | The main causes of climate change are human activities. | Climate change will bring about serious negative consequences. | My local area will be influenced by climate change. | It will be a long time before the consequences of climate change are felt. | Importance of SH | Perceived fairness | Perceived effectiveness | Health professional role | Trust | Expectation | Satisfaction treatment |
| **Demographics** | | | | | | | | | | | | | | | | | | | | | | |
| Age in years | $r_s$ | -- | | | | | | | | | | | | | | | | | | | | |
| Sex/gender | $r_s$ | -,289** | -- | | | | | | | | | | | | | | | | | | | |
| Type of living area, city or rural | $r_s$ | 0,028 | -0,049 | -- | | | | | | | | | | | | | | | | | | |
| Education level in three categories | $r_s$ | -,323** | 0,053 | 0,003 | -- | | | | | | | | | | | | | | | | | |
| Having children | $r_s$ | -,496** | ,152** | -,099* | ,117* | -- | | | | | | | | | | | | | | | | |
| Having grandchildren | $r_s$ | -,684** | ,234** | -0,065 | ,245** | ,546** | -- | | | | | | | | | | | | | | | |

| | | | | | | | | | | | | | | |
|---|---|---|---|---|---|---|---|---|---|---|---|---|---|---|
| Number of GP visits last 12 months | $r_s$ | 0,093 | 0,058 | 0,02 | -,116* | -0,005 | -0,053 | -- | | | | | | |
| Trust in the GP | $r_s$ | 0,002 | -,112* | -0,087 | ,104* | 0,071 | -0,018 | 0,04 | -- | | | | | |
| Self-rated health status | $r_s$ | -,245** | -0,096 | 0,043 | ,208** | ,120* | ,179** | -,318** | 0,054 | -- | | | | |
| **Overall Opinion on Climate Change** | | | | | | | | | | | | | | |
| I believe that climate change is real. | $r_s$ | -,176** | 0,022 | -0,096 | ,271** | ,194** | ,170** | 0,021 | ,128* | 0,07 | -- | | | |
| The main causes of climate change are human activities. | $r_s$ | -,198** | 0,06 | -0,075 | ,259** | ,177** | ,145** | 0,04 | ,111* | ,192** | ,574** | -- | | |
| Climate change will bring about serious negative consequences. | $r_s$ | -,156** | ,103* | -0,08 | ,246** | ,184** | ,141** | 0,045 | 0,092 | 0,081 | ,649** | ,649** | -- | |
| My local area will be influenced by climate change. | $r_s$ | -0,087 | ,107* | -0,052 | ,186** | 0,074 | 0,091 | 0,051 | 0,06 | 0,028 | ,557** | ,486** | ,691** | -- |

| | | | | | | | | | | | | | | | | | | | | | | |
|---|---|---|---|---|---|---|---|---|---|---|---|---|---|---|---|---|---|---|---|---|---|---|
| It will be a long time before the consequences of climate change are felt. | $r_s$ | 0,076 | -,111* | -0,001 | -,139** | -,124* | -0,069 | 0,028 | 0,019 | -0,027 | -,490** | -,363** | -,522** | -,495** | -- | | | | | | | |
| **Constructs in SHQ_GP** | | | | | | | | | | | | | | | | | | | | | | |
| Importance of SH | $r_s$ | 0,066 | ,133** | -0,024 | 0,069 | -0,025 | -0,004 | -0,002 | 0,05 | 0,025 | ,321** | ,228** | ,344** | ,372** | -,338** | -- | | | | | | |
| Perceived fairness | $r_s$ | -0,061 | 0,061 | -0,021 | ,155** | 0,056 | 0,092 | -0,018 | 0,094 | 0,047 | ,341** | ,295** | ,328** | ,322** | -,337** | ,684** | -- | | | | | |
| Perceived effectiveness | $r_s$ | -0,09 | ,118* | -0,035 | ,194** | 0,068 | ,120* | -0,052 | 0,031 | 0,074 | ,388** | ,248** | ,337** | ,396** | -,368** | ,714** | ,719** | -- | | | | |
| Health professional role | $r_s$ | ,197** | -0,03 | 0,008 | 0,004 | -0,05 | -,120* | -0,012 | 0,023 | 0,016 | ,206** | ,149** | ,221** | ,252** | -,186** | ,714** | ,545** | ,535** | -- | | | |
| Trust | $r_s$ | -0,003 | -0,037 | 0,018 | 0,076 | 0,021 | 0,035 | -0,002 | ,124* | ,110* | ,273** | ,217** | ,251** | ,298** | -,274** | ,626** | ,671** | ,616** | ,548** | -- | | |
| Expectation | $r_s$ | 0,095 | 0,009 | 0,025 | 0,051 | -0,013 | -0,008 | -0,041 | ,108* | 0,074 | ,307** | ,216** | ,276** | ,314** | -,293** | ,766** | ,740** | ,732** | ,722** | ,742** | -- | |
| Satisfaction treatment | $r_s$ | -0,073 | 0,045 | -0,026 | 0,071 | 0,052 | 0,055 | 0 | 0,087 | ,130** | ,252** | ,250** | ,286** | ,283** | -,229** | ,595** | ,646** | ,621** | ,473** | ,687** | ,643** | -- |

*Table S3 – Correlation table for demographics, overall opinion on climate change and SHQ_GP constructs using Spearman's' Rho*

** Correlation is significant at the 0.01 level (2-tailed).

* Correlation is significant at the 0.05 level (2-tailed).
